# Genetic correlations between gut microbiota genera and Alzheimer’s Disease

**DOI:** 10.1101/2022.09.26.22280374

**Authors:** Davis Cammann, Yimei Lu, Melika J. Cummings, Mark L. Zhang, Joan Manual Cue, Jenifer Do, Jeffrey Ebersole, Xiangning Chen, Edwin C. Oh, Jeffrey L. Cummings, Jingchun Chen

## Abstract

1.

**Background:** Dysbiosis of the human gut microbiota has been implicated in the development of Alzheimer’s disease (AD). However, the genetic correlation between gut microbiota and AD is not well addressed.

**Methods:** Using the largest genome-wide association study (GWAS) of gut microbiota genera from the MiBioGen consortium, we conducted the “best-fit” model from PRSice-2 to determine the genetic correlation between 119 genera and AD in a discovery sample (case/control: 1,278/1,293); we then replicated our findings in an independent sample (case/control: 799/778) and further performed meta-analyses to confirm the correlation. Finally, we conducted a linear regression to assess the correlation between the PRSs for the significant genera and the *APOE* genotype.

**Results:** In the discovery sample, 20 gut microbiota genera were initially identified genetically associated with AD. Three genera (*Eubacterium fissicatena* as a protective factor, *Collinsella* and *Veillonella* as a risk factor) were validated in the replication sample. Meta-analysis confirmed nine genera to have a significant correlation with AD, three of which were significantly associated with the *APOE* rs429358 risk allele in a direction consistent with their protective/risk designation in AD association. Notably, the proinflammatory genus *Collinsella*, identified as a risk factor for AD, was positively correlated with the *APOE* rs429358 risk allele in both samples.

**Conclusion:** Host genetic factors influencing the abundance of nine genera are significantly associated with AD, suggesting that these genera may serve as biomarkers and targets for AD treatment and intervention. Our results highlight that proinflammatory gut microbiota might promote AD development through interaction with *APOE*. Larger datasets and functional studies are required to understand their causal relationships.

## 2. Introduction

Alzheimer’s disease (AD), the most common form of dementia, is a neurodegenerative disorder characterized by a multitude of pathological and clinical hallmarks such as a progressive decline in cognitive function and the buildup of toxic β-amyloid and tau proteins (1,2). Due to growing elderly populations worldwide, the number of individuals with dementia is projected to reach 150 million globally by the year 2050 (3). Despite this growing burden to world health, the mechanisms underlying the disease pathology are not fully understood, impeding the development of optimally effective treatments (4). Neuroinflammation has emerged as a key feature of AD with mechanistic and treatment implications due to the central role of microglia and inflammation in brain health (5,6). There remains an urgent need to understand the genetic risk factors and pathological basis of neuroinflammation in AD so that individuals with a higher risk can be identified for earlier intervention.

Recently, an association between dysbiosis of the gut microbiome and neuroinflammation has been hypothesized to drive AD. The gut microbiota comprises a complex community of microorganism species that reside in our gastrointestinal ecosystem; alterations in the gut microbiota have been reported to influence not only various gut disorders but also brain disorders such as AD (7,8). The human gut microbiota has been suggested to modulate the brain function and behavior via the microbiota-gut-brain axis (MGBA), a bidirectional communication system connecting neural, immune, endocrine, and metabolic pathways (9). Observational studies across multiple countries show reductions in gut microbiota diversity in AD patients compared to cognitive normal controls (10–12). Current research indicates that bacteria populating the gut microbiota are capable of releasing lipopolysaccharide (LPS) and amyloids, which may induce microglial activation in the brain and contribute to the production of proinflammatory cytokines associated with the pathogenesis of AD (13). The secretion of these biomolecules also harms the integrity of the MGBA and blood-brain-barrier (BBB), which worsens with increasing dysbiosis (8,14). The composition of the human gut microbiota and risk for AD have been suggested as heritable traits (2,15). Apolipoprotein E ε4 (*APOE* ε4), the most well-established risk gene for AD, has recently been shown to correlate with microbiome composition in humans and mouse models of AD (16–18). However, few studies have explored the correlation between *APOE* alleles and microbiome taxa at the human genomic level. In this study, we aim to determine the genetic correlation between the abundance of gut microbial genera and AD diagnosis. We further investigate if gut microbial genera are correlated with *APOE* ε4 alleles.

One promising approach to exploring this relationship is the use of polygenic risk scores (PRS) analyses. PRS is an overall estimate of an individual’s genetic liability for a specific trait. PRSice-2 is designed to calculate the PRS by aggregating and quantifying the effect of many single-nucleotide polymorphisms (SNPs) in the genome weighted by their effect sizes from genome-wide association studies (GWASs) (19). This approach has previously been used to explore the genetic relationship of gut microbial abundance and complex traits like bone mineral density, rheumatoid arthritis, and depression (20–22). In the present study, we used this approach to determine the genetic relationship between 119 microbial genera and AD diagnosis. With the largest GWAS of the human gut microbiota (23), we first conducted PRS analyses in an AD discovery sample to identify the genera genetically correlated with AD. We then verified our results in an independent AD replication sample. Correlation between the top genera and *APOE* genotype was further analyzed by linear regression.

## 3. Methods and materials

### 3.1 Study design overview

We obtained GWAS summary data (n = 18,340) from the MiBioGen consortium initiative (www.mibiogen.org) (23) and requested genotyping data from dbGaP (https://www.ncbi.nlm.nih.gov/gap/). A brief description for these studies is listed in **Table 1**. The overall design of the study is shown in **Figure 1**. Briefly, we used PRSice-2 (19) to calculate PRSs for individuals from our discovery sample. PRSs were calculated based on the summary statistics for 119 microbial genera from the MiBioGen consortium. Significant association between genera and AD diagnosis was considered when the “Best Fit” PRS model with a *p* < 0.00042 (0.05/119 with Bonferroni correction). We then validated the results in an independent sample. We conducted logistic regression analyses between the PRSs of associated genera and AD diagnosis to generate relative odds ratios (OR) for meta-analyses. Multivariate logistic regression was used to determine if the correlation between the PRSs of the associated genera and AD diagnosis was affected by sex, age, and *APOE* genotype. Furthermore, we conducted a linear regression analysis to evaluate the genetic association between the PRSs of nine significant genera and the *APOE* genotype of individuals in our discovery and replication samples. This study was approved by our institutional review board (IRB) at the University of Nevada Las Vegas (UNLV).

**Table 1.**
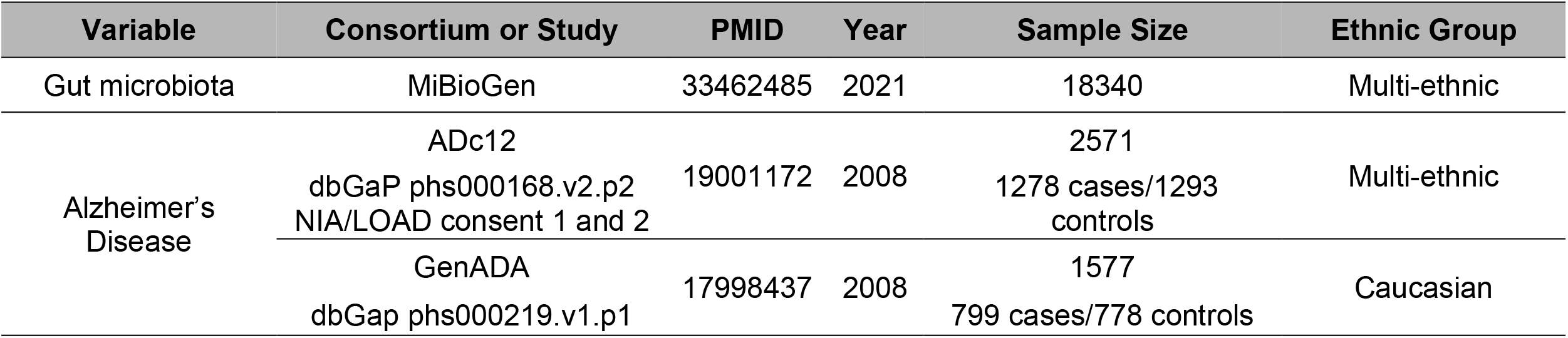
Information for studies used in our analyses.

**Figure 1.**
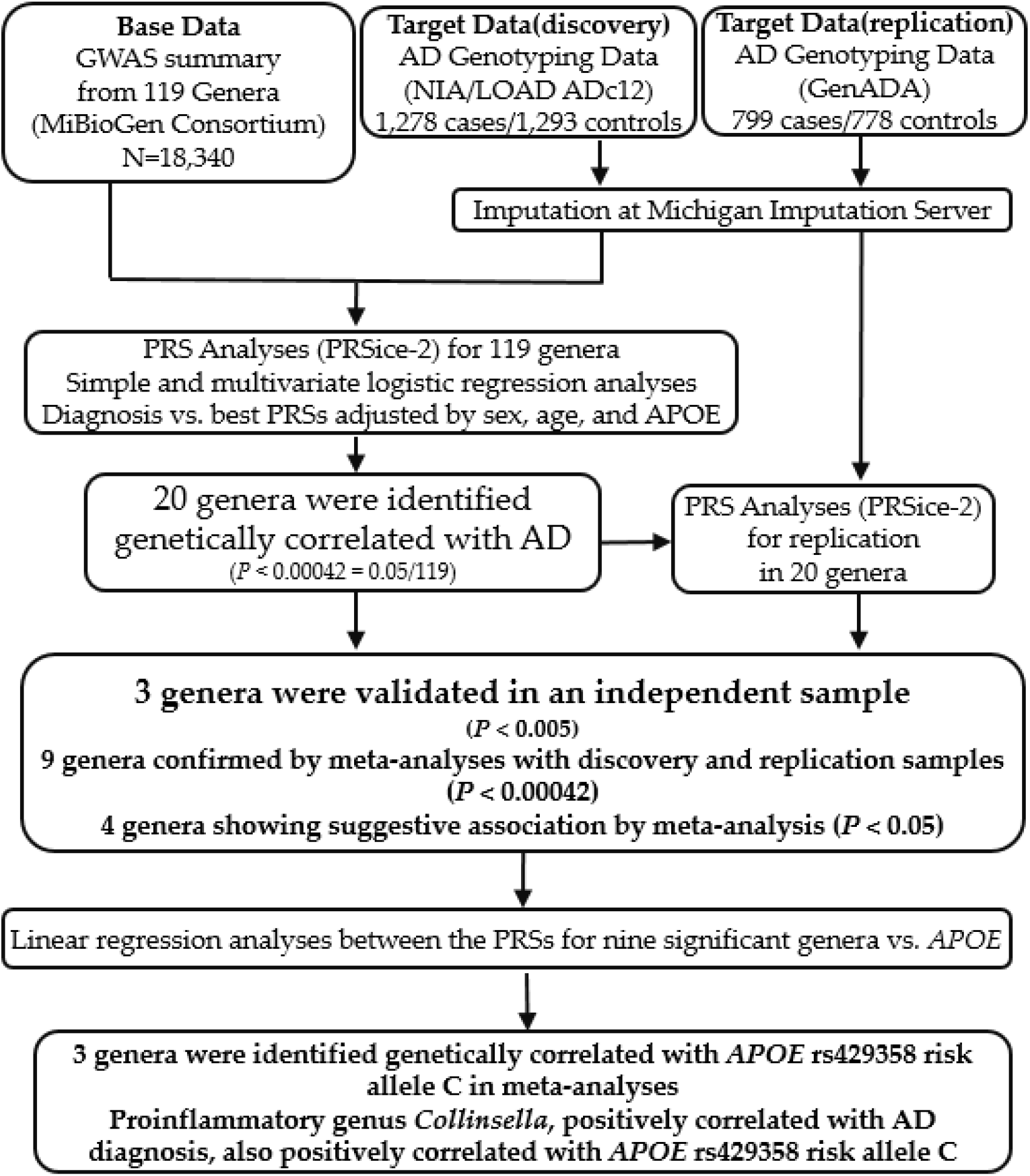
Study design flowchart. In the PRS analysis, “Base” data included summary statistics for 119 microbial genera from the MiBioGen Consortium. “Target” data included AD genotyping data in NIA/LOAD cohort consents 1 and 2 (ADc12) for our discovery sample, and AD genotyping from the GenADA study as our replicate. PRS analyses with PRSice-2, logistic regression analyses, and meta-analyses were used to determine the genetic correlation between the 119 genera and AD diagnosis. 20 genera were found to be significantly genetically associated with AD in the discovery sample, three genera were validated in the replication sample, and nine genera were confirmed by meta-analyses with discovery and replicate samples. Linear regression analyses were used to determine the genetic correlation between the PRSs for nine significant genera and *APOE* genotyping. Three genera were identified genetically correlated with *APOE* rs429358 risk allele C.

### 3.2 Data sources

#### 3.2.1 Microbiome GWAS summary statistics

We used the GWAS summary statistics (base data) of human gut microbiota in the MiBioGen consortium (www.mibiogen.org), which is the largest, multi-ethnic genome-wide meta-analysis of the gut microbiome to date (23). Participants (n = 18,340) were recruited in the original studies, where 16S rRNA gene sequencing profiles from each individual were utilized to characterize the gut microbiota using SILVA as a reference database (24) with truncation of the taxonomic resolution down to the genus level. In total, 31 loci from host genetic variations were associated with gut microbiota taxa abundance at the genome-wide significant threshold (*p* < 5.0 × 10^−8^), most of which (n = 24) were at the genus level (23). We chose the effect sizes of the host SNP-microbiota associations (beta) for our analyses. Beta represents how the host genetic loci affect the relative abundance of each microbiome category (mbQTLs) (23). In the present study, we limited our analyses to the summary statistics from the 119 microbial genera, as 16S rRNA sequencing correlates more accurately with the functional role of gut microbiota at lower taxonomic levels (25).

#### 3.2.2 AD genotyping data (target data): discovery and replication samples

From dbGaP (https://www.ncbi.nlm.nih.gov/gap/), we requested genotyping data from the NIA/LOAD cohort consents 1 and 2 (ADc12) as our discovery sample (26), and the Multi-Site Collaborative Study for Genotype-Phenotype Associations in Alzheimer’s Disease (GenADA) Study as our replication sample (27). In our study, AD cases were considered as any individual with dementia diagnosed with definite, probable or possible AD at any point in their clinical course, according to the Criteria proposed in 1984 by the National Institute of Neurological and Communicative Disorders and Stroke, and the Alzheimer’s Disease and Related Disorders Association (NINCDS-ADRDA) (28). Included controls were neurologically evaluated individuals who were age-matched cognitively normal. Unspecified dementia, unconfirmed controls, and controls with other neurological diseases from the original study were removed from this study, resulting in 1,278/1,293 cases/controls in the discovery sample ADc12, and 799/778 cases/controls in the replication sample GenADA. Demographic characteristics of the ADc12 and Gen/ADA samples are listed in **Table 2**, along with two major *APOE* SNP genotype information. More detailed descriptions of the data can be found in previous studies (26,27,29). In order to maximize genetic variants, we conducted imputation for both discovery and replication samples at the Michigan Imputation Server (minimac4) (https://imputationserver.sph.umich.edu) (30). The 1,000 Genome Phase 3v5 was used as a reference. After the imputation, standard quality control was performed with plink command (--maf 0.01 --hwe 1e-6 --geno 0.01 --mind 0.01) (31,32). The final datasets were composed of 2,571 individuals with 9,997,692 SNPs in the discovery sample, and 1,577 individuals with 8,914,585 SNPs in the replication sample.

**Table 2.**
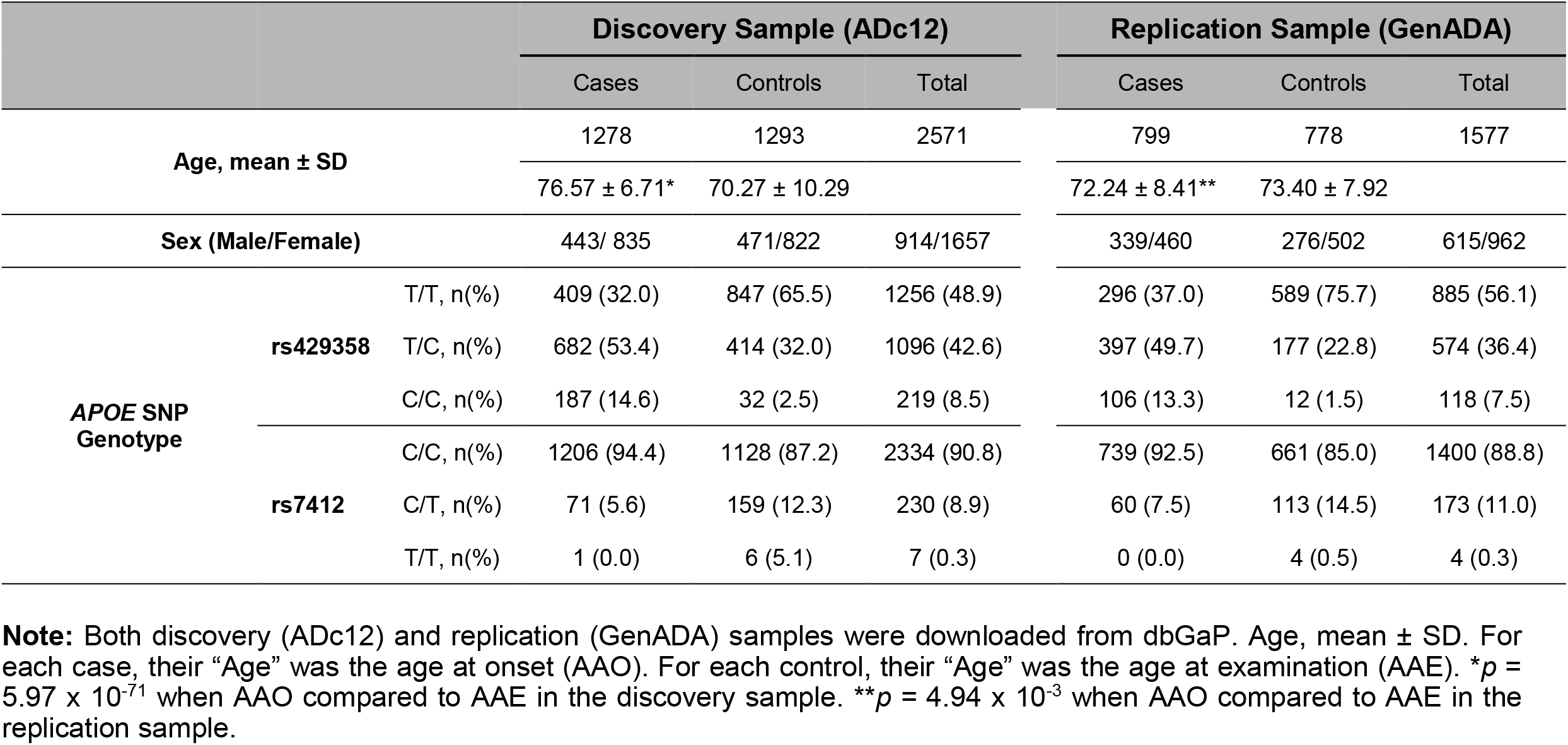
Demographic characteristic of the target data (ADc12 and GenADA) with *APOE* SNP genotyping.

### 3.3 Polygenic Risk Score (PRS) analyses via PRSice-2 software

PRSice-2 software was designed to evaluate the genetic correlation between different traits when provided GWAS summary statistics data from a base trait (base data) and genotyping data from a target trait (target data) (19). PRS is a numerical approximation of an individual’s genetic liability weighed by the allele number and effect size from a set of genetic variants estimated from GWAS summary statistics (33). This approach is called polygenic risk score (PRS) analyses, by which investigators can assess an individual’s genetic risk to a trait of interest and the genetic correlation between different traits (33). As mentioned above, our base GWAS data were from the 119 microbiome genera in the MiBioGen consortium study (23). Our target samples were discovery sample ADc12 (26) and replication sample GenADA (27). In our study, we first calculated PRSs for the 119 microbiome genera in the discovery sample to determine the genetic correlation between the PRSs and AD diagnosis. PRSs for each genus were calculated from the PRSice-2 program for the “best-fit” PRS model. For this purpose, a range of *p* value thresholds was set from 5 × 10^−8^ to 1 with an incremental interval of 0.00005 (--interval 0.00005 --lower 5e-08) with LD clumping (--clump-kb 250kb --clump-p 1.0 --clump-r2 0.1) (19). To validate the significantly associated genera from the discovery sample, we conducted PRS analyses for the significant genera in an independent sample.

To evaluate the overall association from our discovery and replication samples, we rescaled the “best-fit” PRSs of the associated genera with a range of -1 to 1 and performed a simple logistic regression between the rescaled “best-fit” PRSs and AD diagnosis. We then conducted meta-analyses for both samples R packages metafor v3.8-1 (34). A forest plot was generated to visualize the overall AD protective and risk effect across the significant genera using the “forestplot” R package (35). We also created boxplots to compare the PRSs for those significant genera between AD patients and controls in the discovery sample. Box plots were generated by using the R program ggplot2 v.0.4.0 (36). Multivariate logistic regression was conducted by adding sex, age, and *APOE* genotypes (rs429358, rs7412) as covariates.

### 3.4 Linear regression analyses between *APOE* genotyping and PRSs for the nine significant genera

Two *APOE* SNPs, rs429358 minor allele C and rs7412 major allele C, are well-known risk factors for AD (37,38). We performed linear regression analyses to determine the genetic correlation between the PRSs of the nine significant genera and the two *APOE* SNPs. The association was further assessed by linear regression adjusted for sex and age. Boxplots were created using the R packages ggplot2 (v3.3.6), ggpubr (v0.4.0), and stats (v0.1.0) (35,36,39).

### 3.5 Statistical Analyses

The *p* value threshold for significant association in the discovery sample and meta-analyses was set as *p* < 4.20 × 10^−4^ (0.05/119 with Bonferroni correction) (40). For the replication sample, one-side significant level *p* < 0.005 (0.1/20 with Bonferroni correction) was used. For all other statistical analyses, such as linear regression, the ANOVA test, and Wilcoxon signed-rank test, *p* < 0.05 was considered significant. The Wilcoxon signed-rank method was used to test two interrelated samples to see if their sample proportion ranks differed (41). The ANOVA method was utilized to test the association between the PRSs for genera and *APOE* genotype (42).

## 4. Results

### 4.1 PRSs for nine microbiome genera were significantly associated with AD diagnosis

We first calculated the PRSs for the 119 microbiome genera for each individual from the discovery sample ADc12 and conducted the PRS analyses between the PRSs and AD diagnosis using the PRSice-2 program (19). We found that 20 out of the 119 genera were significantly associated with AD diagnosis at the “best-fit” model (*p* < 4.20 × 10^−4^) (**Table 3**). Among these 20 significant genera, six were identified as likely risk genera and 14 potentially protective genera for AD diagnosis. Risk genera included *Alistipes* and *Bacteroides* from the Bacteroidetes phylum, *Lachnospira* and *Veillonella* from the Firmicutes phylum, and *Collinsella* and *Sutterella* from the Actinobacteria and Pseudomonadota phyla, respectively. The most significant risk genus was *Bacteroides* (R^2^ = 0.011, *p* = 3.32 × 10^−6^) at the “best-fit” *p* value of 0.179 with 47,844 SNPs. For protective genera, eleven out of fourteen were from the Firmicutes phylum (*Anaerostipes, Candidatus Soleaferrea, Catenibacterium, Eisenbergiella, Eubacterium coprostanoligenes, Eubacterium fissicatena, Eubacterium nodatum, Intestinibacter, Lachnospiraceae UCG-008, Oscillibacter*, and *Roseburia)*, two were from Actinobacteria (*Adlercreutzia* and *Gordonibacter*), and one was from Bacteroidetes (*Prevotella 9*). The most significant protective genus was *Intestinibacter* (R^2^ = 0.015, *p* = 1.01 × 10^−7^) at the “best-fit” *p* value of 0.190 with 474,833 SNPs.

**Table 3.**
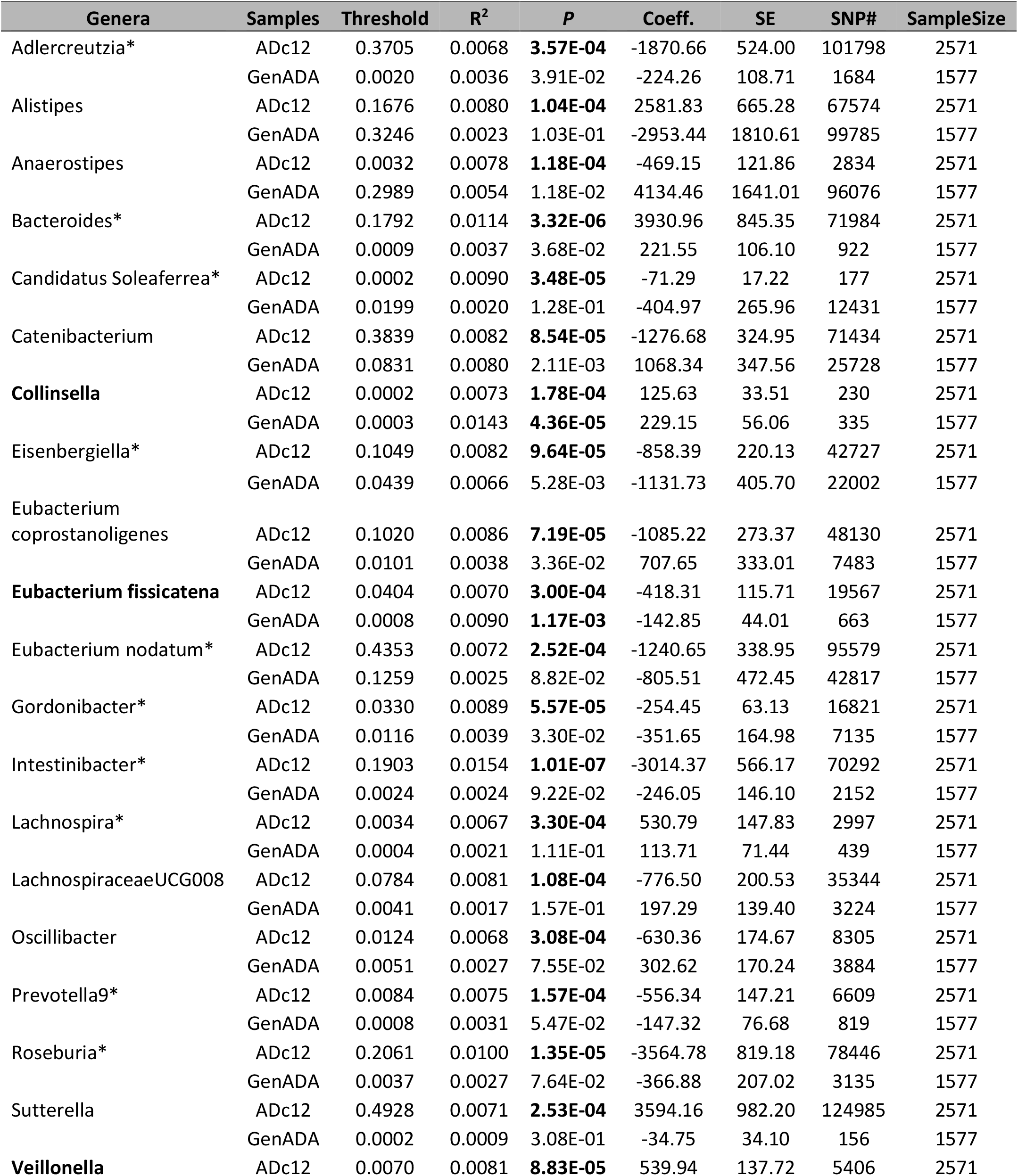
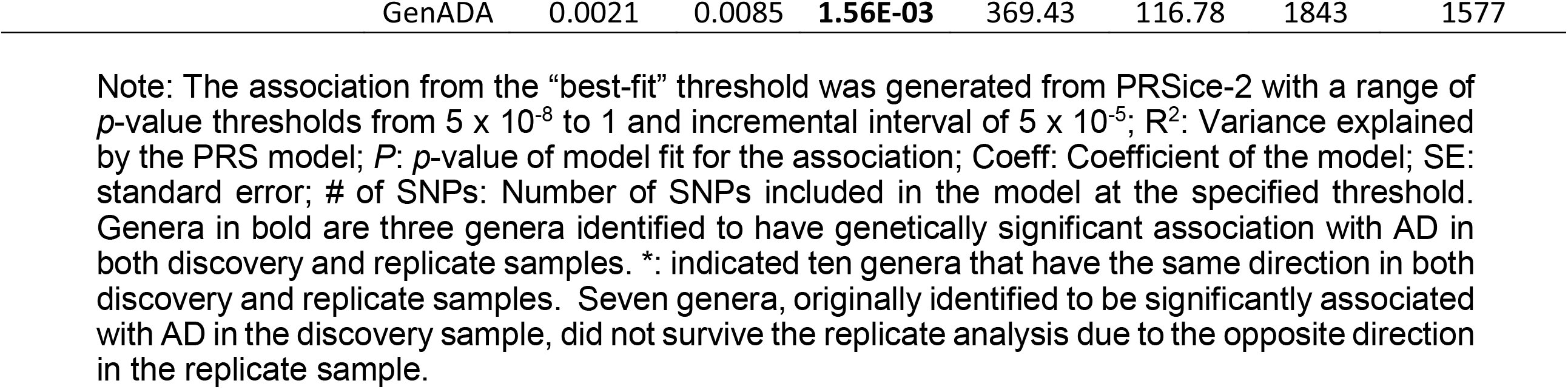
Association between top 20 microbiome genera and AD diagnosis at the “best-fit” model from PRSice-2 in the discovery and replication samples.

To validate our findings for the top 20 genera in the discovery sample, we conducted replication analyses in an independent sample. Three genera, two risk genera (*Collinsella* and *Veillonella*) and one protective genus (*Eubacterium fissicatena*), were successfully validated for their association with AD diagnosis in the replication sample. Ten other genera did not reach the significant *p* value (*p* < 0.005) but had the same effect direction as in the discovery sample. To evaluate the overall association, we conducted meta-analyses with both the discovery and replication samples. As a result, a total of nine genera, including three genera validated from the replication sample, were significantly associated with AD diagnosis (See **Table S1** and **Figure 2**). Six genera—*Adlercreutzia, Candidatus Soleaferrea, Eisenbergiella, Eubacterium fissicatena, Gordonibacter*, and *Prevotella9*—showed as a protective factor for AD, while three genera— *Collinsella, Lachnospira*, and *Veillonella*—showed as a risk factor for AD. From the meta-analyses, *Eisenbergiella* was identified as the strongest protective factor for AD with *p* < 0.0001, and OR = 0.568 (95 % CI 0.450 - 0.716), and *Collinsella* was identified as the strongest risk factor for AD *p* < 0.0001 and OR = 1.869 (95 % CI 1.495 - 2.338). Meta-analyses also found four genera to have a suggestive association (0.00042 < *p* < 0.05) with AD diagnosis genera, of which two genera were a potential risk factor (*Bacteroides* and *Intestinibacter*), and one genus was a protective factor (*Roseburia*). In addition, seven genera—*Alistipes, Anaerostipes, Catenibacterium, Eubacterium coprostanoligenes group, Lachnospiraceae UCG-008, Oscillibacter*, and *Sutterella*—originally identified to be associated with AD, did not show any association due to the opposite effects in the replication sample.

**Figure 2:**
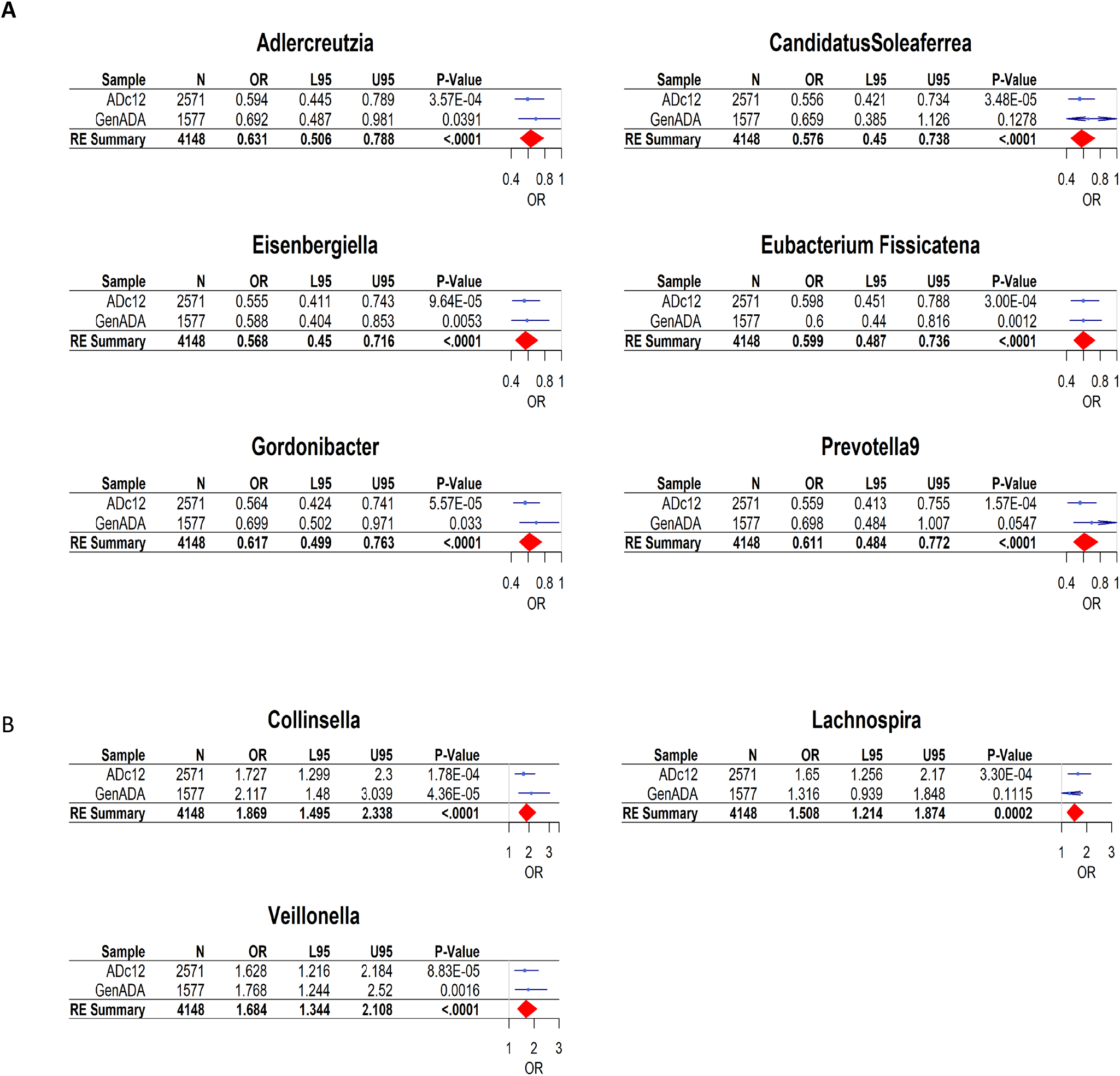
Forest plot of odds ratio and 95% confidence intervals (CI) of rescaled PRSs for nine genera in AD cases compared to controls. The results came from meta-analyses of both discovery (ADc12) and replication samples (GenADA). A. Six genetically predicted genera showed significant association with AD as a protective factor, as *p* < 0.00042 and ORs < 1.0. B. Three genetically predicted genera showed significant association with AD as a risk factor, as *p*-value < 0.00042 and the ORs > 1.0. OR (95%CI): Odds ratio of respective genus with their lower and upper 95 % confidence intervals.

Next, a multivariate logistic regression, including sex, age, and two *APOE* SNPs (rs429358 and rs7412) as covariates, was used to determine any confounding effects on the association between the nine significant genera and AD. As shown in Supplementary **Table S2**, the nine significant genera remained significantly associated with AD diagnosis in the discovery sample (*p* < 0.05), which suggested that the genetic association between PRSs for those genera and AD diagnosis was independent of age, sex, and *APOE*. As expected, age and *APOE* were strongly associated with AD in the multivariate logistic regression analyses. Specifically, age and rs429358 minor allele C were risk factors because of their positive correlation with AD diagnosis, while rs7412 minor allele T was a protective factor due to its negative correlation with AD diagnosis. However, sex did not show any association with AD in our study.

To better visualize the difference of PRSs between AD cases and controls, we constructed a boxplot along with the Wilcoxon signed-rank test (41) for the nine significant genera in the discovery sample. As compared to cognitively normal controls, **Figure 3A** showed that AD patients had lower PRSs for the six likely protective genera (*Adlercreutzia, Candidatus Soleaferrea, Eisenbergiella, Eubacterium fissicatena, Gordonibacter*, and *Prevotella9)*. On the other hand, **Figure 3B** showed AD patients had higher PRSs for the three risk genera (*Collinsella, Lachnospira, and Veillonella*). These results were consistent with the results from our PRS model and logistic regression between PRSs and AD diagnosis.

**Figure 3.**
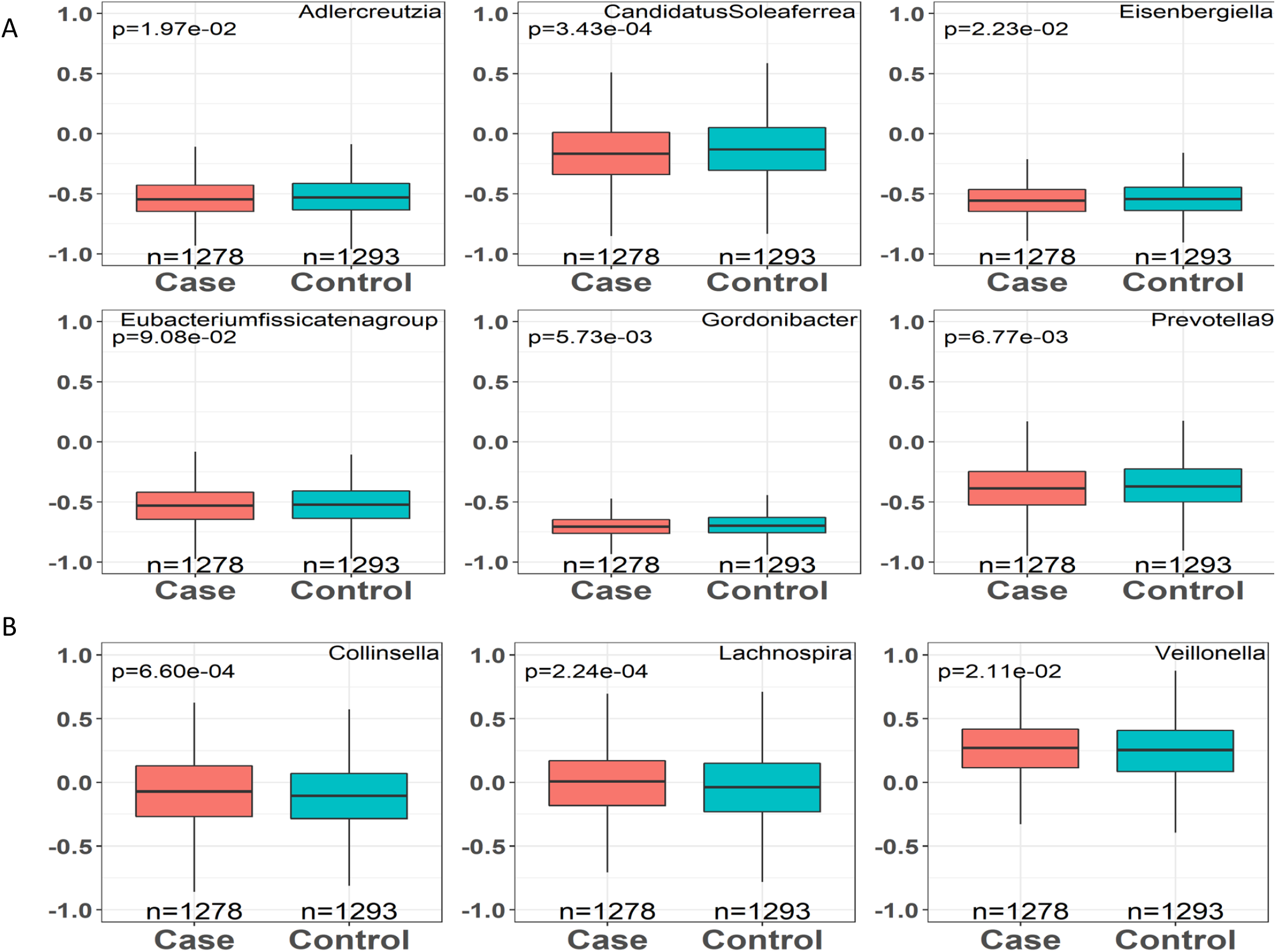
Comparison of rescaled PRSs [-1, 1] for top nine gut genera between AD cases and controls from discovery sample. A. PRSs for six genera were relatively lower as compared AD cases with controls (*p* < 0.05), suggesting those genetically predicted genera were likely to be a protective factor for AD. B. PRSs for three genera were relatively higher as compared in AD cases vs. controls (*p* < 0.05), suggesting those genetically predicted genera were likely be a risk factor for AD. Wilcoxon signed-rank test was applied to generate p values. X-axis: Diagnosis (AD cases/controls) with sample count. Y-axis: rescaled PRSs [-1,1] for each of the top nine genera.

### 4.2 Correlation between PRSs for nine significant genera and *APOE* genotyping

*APOE* is a well-known genetic risk for AD (37,38). Depending on the alleles of two SNPs rs429358 and rs7412, the human *APOE* gene has three major isoforms (ε2, ε3, and ε4) (38). The ε4 isoform is the most influential risk factor beyond age for AD with the presence of a single ε4 allele increasing the risk by 3- to 4-fold compared with ε2 or ε3 allele (37). A few studies have been conducted for the potential links between the *APOE* genotype (rs429358 and rs7412) and the gut microbiota (16–18), but not at the whole genomic level. For this reason, we sought to determine whether there was a genetic link between the PRSs for the nine significant genera and the *APOE* genotype. Linear regression analyses were performed between the rescaled “best-fit” PRSs for the nine significant genera and *APOE* minor alleles at rs429358 and rs7412. *Collinsella* was the only genus that showed significant correlation in both discovery and replication samples. The meta-analyses showed that three out of nine significant genera were correlated with *APOE* rs429358 risk allele C (*p* < 0.05). Notably, *Collinsella*, a proinflammatory genus, was the only genus that was positively correlated with AD and *APOE* rs429358 risk allele C in both discovery and replication samples (*p* < 0.05) (**Table 4**). To illustrate the correlations between PRSs for *Collinsella* and *APOE* rs429358 risk allele C, we constructed a boxplot along with ANOVA analysis (36,43). As shown in **Figure 4**, a positive correlation between PRSs for *Collinsella* and *APOE* rs429358 risk allele C was found in the discovery sample, which indicated that a genetic factor determining *Collinsella* abundance was more likely to occur in individuals with *APOE* rs429358 allele C (CC and TC) as compared to individuals with two T alleles (TT). Overall, our results showed that PRSs for *Collinsella* were significantly higher in AD cases and individuals with APOE risk allele C. Two genera as a protective factor for AD were negatively correlated with *APOE* rs429358 risk allele C (*Adlercreutzia* and *Prevotella9)* (**Table 4**). These associations highlighted that certain members of the microbiome and *APOE* may contribute to modulation in some similar biologic pathways, synergizing in disease risk or protection. The associations between PRSs for the three genera and rs429358 risk allele and were independent of sex and age, as the results remained significant after adjustment for sex and age (Supplementary **Table S2**). For *APOE* rs7412, we did not see any significant correlation.

**Table 4.** Association between PRSs for nine significant gut microbiota genera and *APOE* rs429358.

| Trait | Samples | Coeff. | SE | z-value | p | OR (95%CI) |
| --- | --- | --- | --- | --- | --- | --- |
| **Adlercreutzia** | ADc12 | -0.014 | 0.009 | -1.643 | 0.1006 | 0.99(0.97 - 1.00) |
|  | GenADA | -0.026 | 0.011 | -2.309 | 0.0211 | 0.97(0.95 - 1.00) |
|  | meta-analysis | -0.019 | 0.007 | -2.709 | **0.0068** | 0.98(0.97 - 0.99) |
| Candidatus Soleaferrea | ADc12 | -0.020 | 0.009 | -2.242 | **0.0250** | 0.98(0.96 - 1.00) |
|  | GenADA | -0.004 | 0.007 | -0.486 | 0.6271 | 1.00(0.98 - 1.01) |
|  | meta-analysis | -0.011 | 0.008 | -1.366 | 0.1719 | 0.99(0.97 - 0.99) |
| **Collinsella** | ADc12 | 0.037 | 0.008 | 4.413 | **1.06E-05** | 1.04(1.02 - 1.06) |
|  | GenADA | 0.033 | 0.011 | 2.930 | **0.0034** | 1.03(1.01 - 1.06) |
|  | meta-analysis | 0.036 | 0.007 | 5.278 | **<.0001** | 1.04(1.02 - 1.02) |
| Eisenbergiella | ADc12 | -0.029 | 0.008 | -3.382 | **0.0007** | 0.97(0.96 - 0.99) |
|  | GenADA | 0.002 | 0.011 | 0.180 | 0.8575 | 1.00(0.98 - 1.02) |
|  | meta-analysis | -0.014 | 0.015 | -0.922 | 0.3565 | 0.99(0.96 - 0.96) |
| Eubacterium fissicatena | ADc12 | -0.032 | 0.009 | -3.557 | **0.0004** | 0.97(0.95 - 0.99) |
|  | GenADA | -0.003 | 0.013 | -0.252 | 0.8011 | 1.00(0.97 - 1.02) |
|  | meta-analysis | -0.019 | 0.014 | -1.353 | 0.1762 | 0.98(0.95 - 0.95) |
| Gordonibacter | ADc12 | -0.032 | 0.009 | -3.460 | **0.0005** | 0.97(0.95 - 0.99) |
|  | GenADA | -0.004 | 0.012 | -0.352 | 0.7248 | 1.00(0.97 - 1.02) |
|  | meta-analysis | -0.019 | 0.014 | -1.398 | 0.1621 | 0.98(0.95 - 0.95) |
| Lachnospira | ADc12 | 0.014 | 0.009 | 1.524 | 0.1277 | 1.01(1.00 - 1.03) |
|  | GenADA | 0.014 | 0.012 | 1.169 | 0.2425 | 1.01(0.99 - 1.04) |
|  | meta-analysis | -0.017 | 0.018 | -0.969 | 0.3324 | 0.98(0.95 - 0.95) |
| **Prevotella9** | ADc12 | -0.012 | 0.008 | -1.486 | 0.1373 | 0.99(0.97 - 1.00) |
|  | GenADA | -0.020 | 0.011 | -1.836 | 0.0666 | 0.98(0.96 - 1.00) |
|  | meta-analysis | -0.015 | 0.007 | -2.291 | **0.0220** | 0.99(0.97 - 0.99) |
| Veillonella | ADc12 | 0.024 | 0.008 | 2.946 | **0.0032** | 1.02(1.01 - 1.04) |
|  | GenADA | -0.008 | 0.011 | -0.664 | 0.5071 | 0.99(0.97 - 1.01) |
|  | meta-analysis | 0.0094 | 0.016 | 0.5905 | 0.5549 | 1.01(0.98 - 1.04) |

Note: Genera in bold are three genera identified to have genetically significant correlation with *APOE* rs429358 minor allele C in the meta-analysis ( $p < 0.05$ ). *Collinsella* was the only genus that showed significant correlation in both discovery and replication samples.

**Figure 4.**
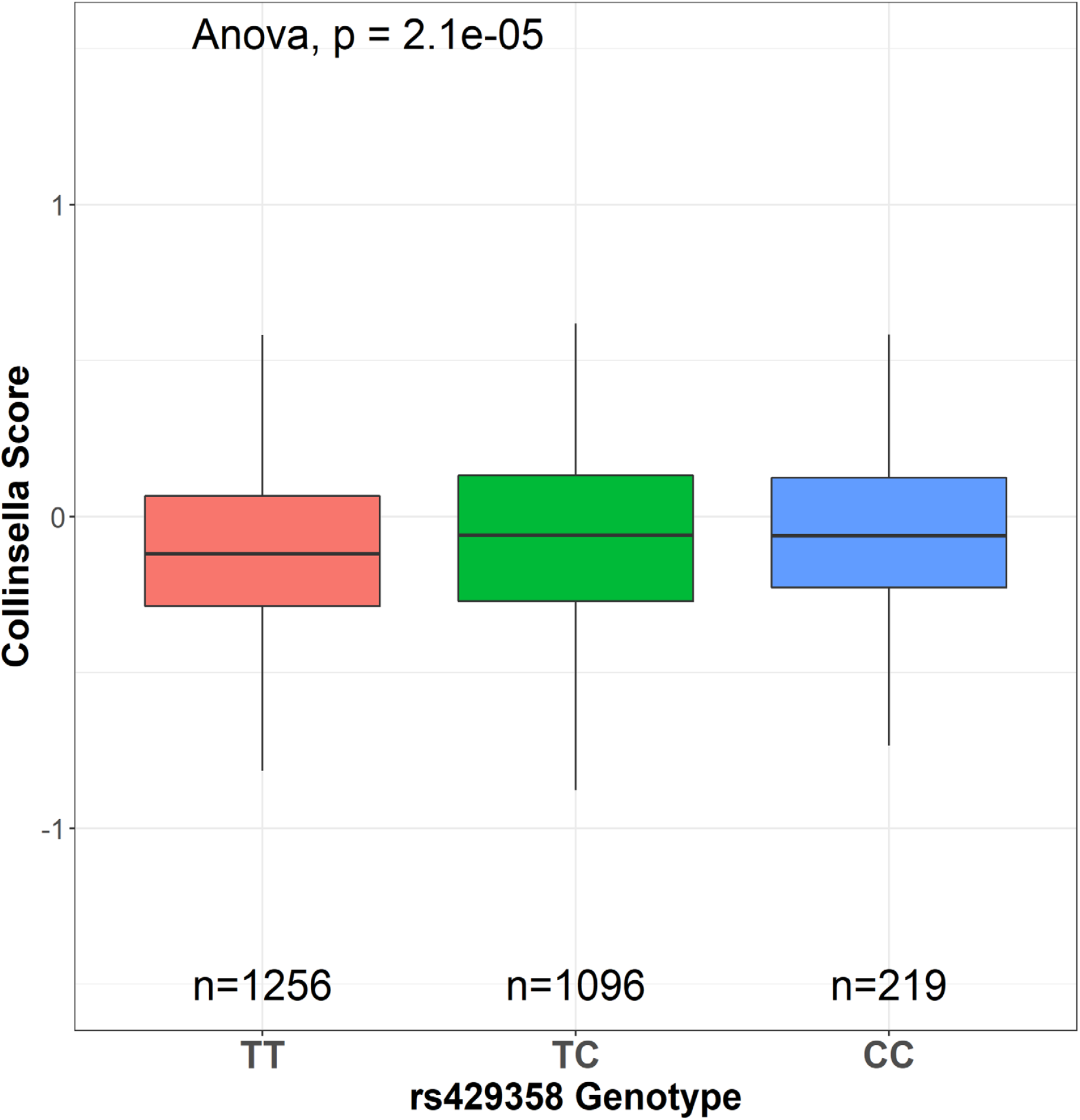
Genetic association between PRSs for *Collinsella* and *APOE* rs429358 minor allele C in the discovery sample. Proinflammatory genus *Collinsella* that was positively associated with AD showed a positive correlation with *APOE* rs429358 minor allele C.

### 4.3 Association between microbiome abundance and *APOE* genotype

To further investigate *APOE* genotype association with the abundance of all the gut microbiota genera, we retrieved summary statistics for the two *APOE* SNPs rs429358 and rs7412 directly from the 119 genera GWAS summary statistics in the MiBioGen consortium study. As shown in **Table 5**, rs429358 was marginally correlated with the abundance of nine genera, and rs7412 was marginally associated with the abundance of eight genera (*p* < 0.05). Together, these findings indicate that the *APOE* genotype may have some impact on the microbiome abundance at the genus level, and that the association may synergistically contribute to the risk for human diseases such as AD. Our results open the door for future studies to explore the role of the interaction between *APOE* and the gut microbiota and find a new target for treatment in human diseases.

**Table 5.** List of gut microbiome genera that were nominally associated with *APOE* SNPs rs429358 and rs7412.

| rsID (CHR:BP:Effect Allele) | Microbiome Genera | Beta | SE | SZ | P | N | # cohorts |
| --- | --- | --- | --- | --- | --- | --- | --- |
| rs429358 (19:45411941:C) | Bacteroides | -0.044 | 0.015 | -2.881 | 0.0040 | 18173 | 23 |
|  | Butyricimonas | -0.044 | 0.020 | -2.241 | 0.0250 | 10657 | 23 |
|  | Dorea | 0.030 | 0.015 | 2.127 | 0.0334 | 17494 | 23 |
|  | Eubacterium coprostanoligenes | 0.033 | 0.015 | 2.120 | 0.0340 | 17261 | 23 |
|  | Faecalibacterium | -0.035 | 0.015 | -2.188 | 0.0287 | 17960 | 23 |
|  | Olsenella | 0.064 | 0.033 | 1.993 | 0.0462 | 3721 | 13 |
|  | Parasutterella | 0.040 | 0.019 | 2.216 | 0.0267 | 11291 | 23 |
|  | Senegalimassilia | 0.050 | 0.024 | 2.207 | 0.0273 | 6888 | 21 |
|  | Veillonella | 0.044 | 0.021 | 2.032 | 0.0421 | 9194 | 23 |
| rs7412 (19:45412079:T) | Butyricicoccus | 0.048 | 0.021 | 2.468 | 0.0136 | 15637 | 20 |
|  | Collinsella | 0.052 | 0.023 | 2.214 | 0.0268 | 12811 | 19 |
|  | Coprococcus3 | 0.050 | 0.022 | 2.344 | 0.0191 | 14323 | 20 |
|  | Eubacterium hallii | 0.053 | 0.021 | 2.453 | 0.0142 | 14846 | 20 |
|  | Lachnospiraceae UCG001 | -0.071 | 0.027 | -2.534 | 0.0113 | 9357 | 20 |
|  | Olsenella | -0.103 | 0.043 | -2.346 | 0.0190 | 3721 | 13 |
|  | Ruminococcaceae UCG004 | 0.080 | 0.028 | 2.901 | 0.0037 | 9049 | 20 |
|  | Senegalimassilia | 0.082 | 0.032 | 2.584 | 0.0098 | 6508 | 18 |

Note: The data were extracted from the largest GWAS of human gut microbiome conducted among 24 multi-ethnic cohorts in the MiBioGen consortium (www.mibiogen.org) (24). rsID (CHR:BP:Effect Allele): the rs-number of single nucleotide polymorphisms, chromosome number (CHR), base pair (BP), with the effect allele (genome assembly GRCh37/hg19). Microbiome: only contains the genus level with significant abundance associated with *APOE* two SNPs. Beta: Beta coefficient. SE: Standard Error. SZ: Weighted sum of z-scores. P: p-value (p<0.05 was considered as nominal association between the microbiome genera and *APOE*). N: sample count. # Cohorts: Number of cohorts involved.

## 5. Discussion

The microbiota is a complex ecosystem that comprises more than 100 trillion symbiotic microbial cells in the human body, of which 95 % inhabit the human gut (44). The bacteria belonging to phylum Firmicutes and Bacteroidetes form a significant proportion (90 %) of the adult gut microbiota, and Actinobacteria composes the rest (45). Recently, significant evidence has shown that the gut microbiota influences normal systemic physiological homeostasis, and that dysbiosis of gut microbiota may contribute to the pathogenesis of brain diseases, including AD. The gut microbiota interacts with the central nervous system (CNS) across the MGBA via microbial components, metabolic products, and neural stimulation. In this study, we leveraged the extensive GWAS data to study the genetic correlation between gut microbiota genera and AD diagnosis. PRSs for 20 genera were initially found significantly associated with AD in the discovery sample, three of which were validated in the independent replication study. Further meta-analyses from our discovery and replication samples identified a strong genetic association between nine gut microbiota genera and AD diagnosis. Six genera were negatively associated with AD diagnosis and three genera were positively correlated with AD diagnosis. The negative association means that the abundance of these genera is negatively associated with AD diagnosis, thus, PRSs for such genera could be seen as a protective factor for the disease. Similarly, positive association means that the abundance of those genera is positively associated with AD diagnosis, thus, PRSs for these genera would be seen as a risk factor for the disease. Genera identified as a protective factor were primarily from the Firmicutes phylum (*Candidatus Soleaferrea, Eisenbergiella*, and *Eubacterium fissicatena)* as well as from Actinobacteria (*Adlercreutzia, Gordonibacter*) and Bacteroidetes (*Prevotella9*). Positively correlated, or risk-associated genera were from phyla including Firmicutes (*Lachnospira* and *Veillonella*) and Actinobacteria (*Collinsella*).

In the discovery sample, the correlation of the nine significant genera remained statistically significant after being adjusted for sex, age, and two *APOE* SNPs (rs429358 and rs7412), suggesting that the genetic correlation between the genera and AD diagnosis was independent of age, sex, or *APOE* genotype. In addition, we found that three of the nine significant genera showed a strong correlation with the *APOE* rs429358 risk allele C via linear regression analyses. Interestingly but not surprisingly, the genera showing positive correlation with *APOE* rs429358 risk allele C tend to have a positive (risk) association with AD, while the genera showing negative correlation with *APOE* rs429358 risk allele C have a negative (protective) association with AD.

In our analyses, *Collinsella* from the phylum Actinobacteria was found to be a risk factor for AD in the discovery samples and then was validated in replication sample. *Collinsella* was also found to be positively correlated with *APOE* rs429358 risk allele C in both samples. *Collinsella* is a proinflammatory genus that is associated with cumulative inflammatory burden in rheumatoid arthritis patients (46). Studies also found that *Collinsella* is abundant in atherosclerosis and type-2 diabetes patients (47,48). Increased abundance of this genus has been observed in AD transgenic mice and AD patients (49,50). As type-2 diabetes is a well-established risk factor for AD (51), our association of genetically-predicted *Collinsella* with AD cases further supports a connection between the two diseases. The strong association between *Collinsella* and AD diagnosis along with its strong correlation with the *APOE* rs429358 risk allele C may provide new insight into how the interaction between the *APOE* ε4 gene and the AD-risk pathogens promotes the risk for AD development. Targeting the risk microbiota might be an effective therapeutic strategy for AD patients with *APOE* ε4 allele. Further studies to evaluate the impact of the *APOE*-associated microbiota on AD-related phenotypes will be necessary to determine if alterations in the gut microbiome represent a novel mechanism and target for the development of potentially novel interventions for AD.

Three genera of the Firmicutes phylum, *Candidatus Soleaferrea, Eisenbergiella*, and *Eubacterium fissicatena group*, had a negative association with AD diagnosis. *Eisenbergiella* and *Eubacterium fissicatena group* are known to contain species that metabolize the short-chain fatty acid (SCFA) butyrate from dietary carbohydrates (52–54). Butyrate is a major SCFA metabolite in the colon and has been shown to be a critical mediator of the colonic inflammatory response. Alongside its anti-inflammatory properties, butyrate is also essential in maintaining the permeability of the gut across the MGBA (55,56). To our knowledge, there are no studies that presently implicate these genera in neurodegenerative diseases. Nevertheless, *Eubacterium fissicatena*, one of the three genera that was validated in the replication sample was found to decrease in mice with periodontitis, a disease linked to a prediabetes condition. Within the limitations of their study, the authors claimed that the gut microbiota may mediate the influence of periodontitis on prediabetes (57). On the other hand, studies found that *Candidatus Soleaferrea* tends to have a higher abundance in controls as compared to a rat prediabetic condition (58). Interestingly, the study found that colon Glucagon-like peptide-2 (GLP-2) was positively correlated with an abundance of *Candidatus Soleaferrea* (58). GLP-2, secreted from the metabolites (e.g., SCFA) of gut microbiota, is typically considered a trophic hormone involved in maintaining intestinal epithelial morphology and function. GLP-2, along with GLP-1, has been shown to possess neuroprotective properties in AD animal models (59,60). It is not clear exactly why *Eubacterium fissicatena* and *Candidatus Soleaferrea* have a protective effect against AD; however, based on the literature, both seem to have a higher abundance in the controls compared to prediabetes-related condition, which may provide some clue for future study on their AD protection. Further study would provide more insights into how to utilize the microbiome and their metabolic molecules to improve AD treatment.

In addition, we identified two Firmicutes genera as a risk factor for AD (*Lachnospira* and *Veillonella*). The association with *Veillonella* was validatedin the replication sample. *Veillonella* is a normal bacterium in the intestines and oral mucosa of mammals. Consistently, other studies found that AD patients seem to have more abundance of *Veillonella* in their oral microbiome (61). Lately, another study found that overabundance of *Veillonella parvula* (belonging to *Veillonella* genus) promotes intestinal inflammation by activating macrophages via the lipopolysaccharide-Toll-like receptor 4 (LPS-TLR4) pathway (62). Increasing intestinal inflammation by *Veillonella* might also promote AD through MGBA. On the other hand, gut *Lachnospira* and *Veillonella* species have also been identified as beneficial or commensal to gut health, such as *Lachnospira* being protective against Crohn’s disease, or *Veillonella* interacting with *Streptococcus* species to modulate immune responses in the small intestine (63,64). In an observational study from a Chinese group, patients with AD had decreased *Lachnospira* at the genus level compared with healthy controls (65).

*Prevotella9*, belonging to Bacteroidetes, was identified as a protective factor against AD in our study. There is an inconsistent relationship between *Prevotella* abundance and intestinal diseases in human studies (66). In humans, *Prevotella* is more common in populations with plant-based and high-carbohydrate diets (67). Conversely, *Bacteroides* is more abundant in those consuming “western” diets high in protein and fat (68). One study found that a higher *Prevotella*-*Bacteroides* ratio in the gut was predictive of the effectiveness of a 24-week weight-loss regimen in obese patients (69). Another major study showed that *Prevotella* was higher in individuals with greater adherence to Mediterranean diets, which is thought to be protective against neurodegenerative diseases (70–72). The “protective” effects of *Prevotella* abundance may come from the positive dietary effects on the genus. In addition, *Prevotella* was also found to be negatively correlated to *APOE* rs429358 risk allele. It is not clear how the *APOE* gene interacts with the microbiome to provide the protective effect. Nevertheless, more work needs to be done to understand the association between the gut microbiota and *APOE*.

Two protective genera, *Gordonibacter* and *Adlercreutzia*, are from the Actinobacteria phylum. These genera tend to produce metabolites beneficial to mitochondrial function, namely Urolithin-A (UA) and Equol (73,74). UA is an anti-inflammatory compound that enhances mitophagy, the removal of dysfunctional mitochondria in a cell (75). Impaired mitophagy is part of the pathogenesis of AD, as well as general aging processes, making UA and *Gordonibacter* species promising targets for therapeutics against the disease (76). Equol is an estrogen-like compound that reduces microglial inflammation when stimulated by LPS and downregulates genes in neurons related to apoptosis (77). The positive effects of these bacterial metabolites likely drive the protective association of *Gordonibacter* and *Adlercreutzia* abundance with AD that we found in this study.

The strengths of our study include the use of the largest available GWAS of gut microbiota taxa to date, the identification of multiple genera genetically associated with AD in the discovery, and replication samples after a strict Bonferroni correction. The use of logistic regression alongside our initial PRS analyses allows us to adjust for potential confounders such as sex, age, and *APOE* alleles and further validate that the association was independent of those confounders. Additionally, we are the first to study the genetic correlation between the gut microbiota and *APOE* gene at the human genomic level. We found that three genera were associated with the *APOE* risk allele at rs429358, one of the strongest genetic risk variants for AD. The identified genetic relationship may provide candidate bacteria for subsequent causal relationships and functional studies.

However, there are several limitations to our study. First, the sample size for the microbiome GWAS may still not be large enough to truly cover the effect size of the host genetic variants, even though the MiBioGen study has the largest sample size compared with other microbiome GWASs. The sample size for our target genotyping was also relatively small. Because of this, we may not have enough power to detect some of the associations in our meta-analyses. For example, two genera, *Bacteroides* and *Intestinibacter*, were originally identified to be the strongest risk and protective factors, respectively, from the discovery sample, however, meta-analyses only found a suggestive association with AD. The modest sample size might be one of the reasons. Heterogeneity between the discovery and replication samples could be another reason. Studies have found that *Bacteroide*s is capable of secreting LPS as an endotoxic biomolecule, which has been implicated in pathological endothelial dysfunction of the gut and can induce neuroinflammation in microglia cells (78–80). For *Intestinibacter*, its protective role for AD is unknown; however, the reduction of *Intestinibacter* is a well-known microbial biomarker of Metformin-induced gut dysbiosis in the treatment of type-2 diabetes (81–83). Future studies with larger sample sizes would be more capable of drawing solid conclusions about the genetic connection between the gut microbiota and AD. Second, our genotyping data was specifically drawn from individuals in the United States, which limits the generalization of our conclusions when applied to other nations or ethnic groups. More diverse genotyping and GWAS datasets would enable us to capture the variability in risk for AD across different ethnicities. Third, the 16S rRNA sequencing used to generate genetic associations in the “base” GWAS only provides taxa resolution to the genus level. Understanding the role of bacterial species that may drive the pathology of AD will require more studies of specific species.

Overall, our novel findings provide new insights into understanding AD and point to new therapeutic targets for earlier intervention in the disease. Further investigations are needed to explore the microbial alterations in larger cohorts of AD patients from different ethnic backgrounds and identify the causal effect between the gut microbiome and AD.

## Data Availability

All data produced in the present study are available upon reasonable request to the authors

## 6. Supplementary Materials

### Supplementary Tables

**Table S1**. Logistic regression and meta-analyses between the PRSs for the top 20 gut microbiota genera and AD diagnosis in discovery and replication samples.

**Table S2**. Association between the PRSs for the nine significant gut microbiota genera and AD adjusted for sex, age, and *APOE* genotype in the discovery sample.

**Table S3**. Linear regression and meta-analyses between the PRSs for the nine significant gut microbiota genera and *APOE* rs429358 risk allele C adjusted for sex and age.

## 7. Data availability Statement

Full GWAS summary statistics for mbQTLs are available at the www.mibiogen.org website built using the MOLGENIS framework (99).

## 8. Ethical Approval and Consent to Participate

Contributing studies received ethical approval from their respective institutional review boards (IRB). This study was approved by the IRB at the University of Nevada-Las Vegas (IRB #00002305, 10/12/2021).

## 9. Acknowledgements

We thank the patients for their participation in the MiBioGen consortium, NIA/LOAD ADc12 Cohort, and GenADA studies, and the original investigators who conducted these studies and made the data available.

## 10. Author contribution

J.C. was responsible for the conception, sample request, data analyses, and writing the manuscript. D.C., Y.L., and M.J.C. were responsible for data analyses and writing the manuscript. M.L.Z., J.M.C, and J.D. were responsible for data analyses. X.C., E.O., J.L.C, and J.E. were responsible for writing, critiquing, and revising the manuscript. All authors have read and agreed to the content of the manuscript before sending to journal for publication.

## 11. Conflicts of Interest

The authors declare no conflict of interest.

## 12. Funding

This research was funded to J.C. in part by NIH grant P20GM121325, NIH grant P20GM121325-02S1 and NIH grant P20GM121325_03S2.

## Notes

### Competing Interest Statement

The authors have declared no competing interest.

### Funding Statement

This study was funded by NIH grant P20GM121325, NIH grant P20GM121325-02S1 and NIH grant P20GM121325_03S2

### Author Declarations

IRB of the University of Nevada Las Vegas gave ethical approval for this work

